# How an election can be safely planned and conducted during a pandemic: Decision support based on a discrete event model

**DOI:** 10.1101/2021.07.17.21260634

**Authors:** Nadine Weibrecht, Matthias Rößler, Martin Bicher, Štefan Emrich, Günther Zauner, Niki Popper

**Affiliations:** Institute for Information Systems Engineering, TU Wien, Vienna, Austria; dwh GmbH, dwh simulation services, Vienna, Austria; Association for Decision Support for Health Policy and Planning (DEXHELPP), Vienna, Austria; Trnava University, Faculty of Health Sciences and Social Work, Department of Public Health, Trnava, Slovakia

## Abstract

In 2020, the ongoing COVID-19 pandemic caused major limitations for any aspect of social life and in specific for all events that require a gathering of people. While most events of this kind can be postponed or cancelled, democratic elections are key elements of any democratic regime and should be upheld if at all possible. Consequently, proper planning is required to establish the highest possible level of safety to both voters and scrutineers. In this paper, we present the novel and innovative way how the municipal council and district council elections in Vienna were planned and conducted using an discrete event simulation model. Key target of this process was to avoid queues in front of polling stations to reduce the risk of related infection clusters. In cooperation with a hygiene expert, we defined necessary precautions that should be met during the election in order to avoid the spread of COVID-19. In a next step, a simulation model was established and parametrized and validated using data from previous elections. Furthermore, the planned conditions were simulated to see whether excessive queues in front of any polling stations could form, as these could on the one hand act as an infection herd, and on the other hand, turn voters away. Our simulation identified some polling stations where long queues could emerge. However, splitting up these electoral branches resulted in a smooth election across all of Vienna. Looking back, the election did not lead to a significant increase of COVID-19 incidences. Therefore, it can be concluded that careful planning led to a safe election, despite the pandemic.

**Author summary:** *Why was this study done?:*

- The ongoing COVID-19-Pandemic poses a risk for elections, as these could lead to another spread of the disease.
- Additional hygiene measures, which are inevitable if the election is conducted, could lead to long queues in front of the polling stations on election day.

*What did the researchers do and find?:*

- We describe the defined additional hygiene measures.
- We modeled the whole election process, including the new hygiene concept, to show where extensive queues could form on the election day.
- We show that thanks to the additional hygiene measures, the election actually did not cause a temporary upswing of the epidemic.

*What do these findings mean?:*

- Thanks to careful planning, it is possible to safely hold an election during such a pandemic.
- Our simulation contributed to a safe and smooth election.
- However, it should be stated, that these findings hold for elections in Austria, where long queues in front of polling stations are comparably rare and short.

## Introduction

In 2020, the COVID-19-Pandemic affected almost every area of daily life, including elections. On the one hand, elections necessarily cause a gathering of people, on the other hand, elections are an essential part of vital democratic societies and are not easily canceled or postponed. Consequently, proper planning is required to enable the election process, provide the highest possible level of safety to all participants, and to avoid that the election stokes the spread of the disease.

In Vienna, the municipal council and district council elections were scheduled for October 11^*th*^ 2020. By this time, SARS-CoV-2 case numbers in Austria, especially in Vienna, were on the rise and the risk of infections was high [1]. So the planning committee consulted modeling and simulation experts to help them increasing the level of safety to voters and scrutineers. The key question of the planning process was how to avoid the development of long waiting queues in front of the polling stations, since these may potentially lead to infection clusters.

It must be stated, that, compared to other countries, waiting queues in front of polling stations are usually comparably rare and short in Austria. Unfortunately, to the authors’ knowledge, there is no data on polling queues as of now. Nevertheless, in 2017, when national elections took place in Austria, an article in the “Wiener Zeitung” (“Vienna newspaper”) stated that “there was even a short wait in front of the cabins at 11 a.m.”, indicating that usually there are hardly any waiting queues in Austria [2].

This is a desirable condition which should be preserved, in particular thinking about infection risks in queues. Yet, since additional hygienic measures, which might slow down the voting process, were required for the election in October, election planners were afraid that it might be different this time.

To ensure that the hygiene measures don’t cause long waiting queues, a discrete event simulation (DES) model was developed to simulate the voting process including the hygiene concept in each electoral branch. Thus, possible problematic districts could be identified, so that additional measures like additional voting booths for these districts could be applied.

In this work we show how this DES model contributes to enabling safe election processes in times of epidemics. The primary objective of this work is to describe the model in detail, to explain its parametrization and validation process and to describe how it detects potential bottlenecks in an election process with respect to emergence of long queues. As a secondary objective, we reevaluate the municipal council and district council elections in Vienna in Fall 2020 which have been planned using the simulation tool and identify the benefits of the usage of the model. The aim of this work is to demonstrate how modelling and simulation can help optimizing election processes with respect to avoiding queues and consequently also avoiding infection clusters in epidemic situations. The paper is intended to report a full country-level experience with a particular event, namely the municipal council and district council elections in Vienna. Yet, we consider it of interest for all democracies, especially facing the COVID-19 pandemic.

## Materials and methods

This section describes the COVID-related methodological background, followed by a description of the boundary conditions and a comparison to an already developed agent-based COVID-19 epidemic model. Finally, the last subsection deals with the details of the discrete event model.

Based on early works on agent-based modelling [3], we described the importance of the detection of temporal and spatial evolution of epidemics. The effectiveness of the method for decision support was described in [4]. In January 2020 our group developed an agent-based COVID-19 epidemic model [5, 6], which is still in heavy use for counseling of political decision makers in Austria [7]. The present question revealed an interesting new chance for us. While the epidemic model concentrates on the overall spread of the disease throughout the Austrian population, it models the places where infections take place, like households, schools, work places, or leisure facilities as chance encounters. This is of course necessary due to the much larger population size. The COVID-19 model analyses the spread of the disease for the whole Austrian population, and therefore uses nearly 9 million agents. In contrast, there were about 1.4 million people eligible to vote at the present elections, from which we expected 30 - 60 % to turn up at polling stations and the rest to use postal votes or not to vote at all.

Moreover, the COVID-19 model investigates a much longer time frame. It predicts the spread of the virus over a time frame ranging from several weeks to months, while the election takes place over the course of 10 hours. The concise character of the presented problem enabled us to model the election process in more detail.

### Additional hygiene measures due to COVID-19

Vienna consists of 23 districts, which are split up into 1456 voting districts (at the time of the elections 2020). Each voting district contains a polling station with an average number of 937 persons being entitled to vote. [8] These voting districts are visualized in Figure 1, colored by the number of persons entitled to vote.

**Fig 1.**
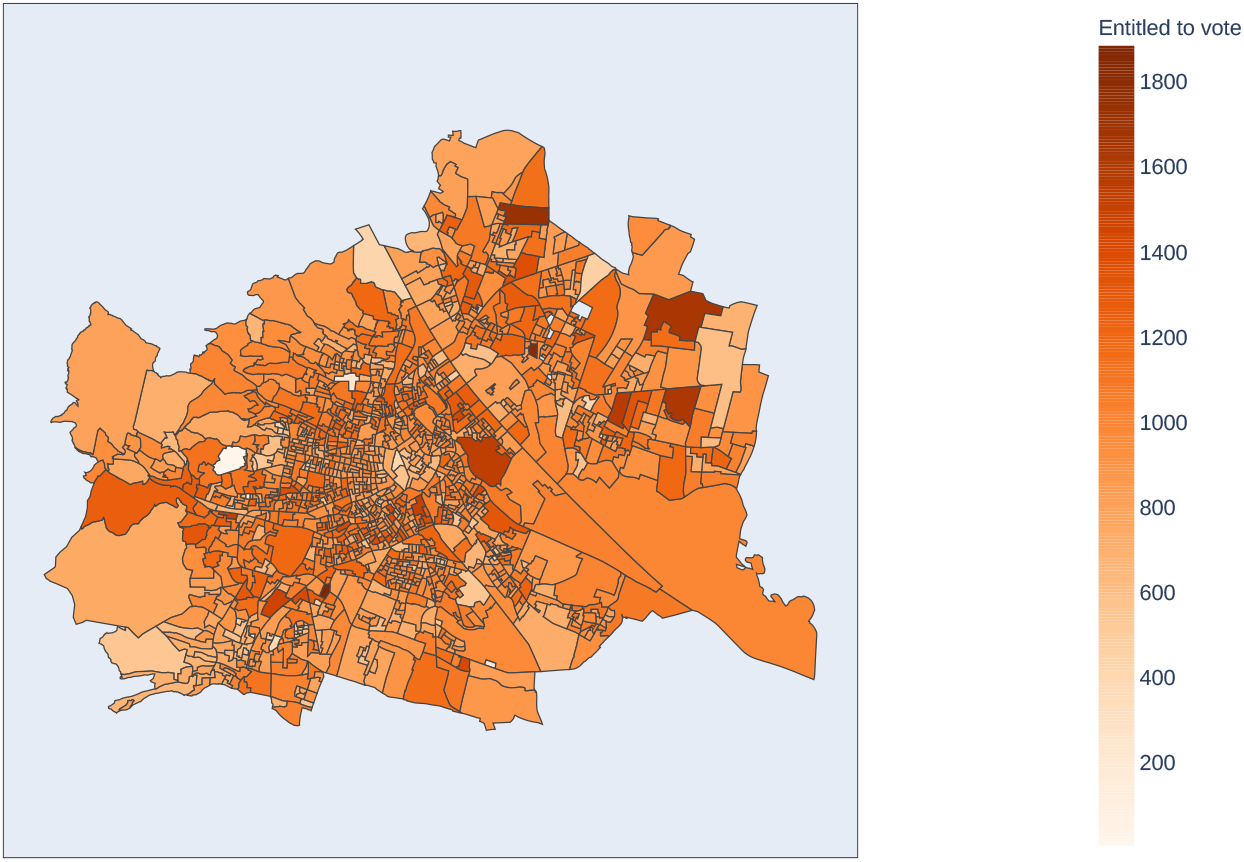
Map of Vienna, indicating the polling districts and the number of people entitled to vote in each district. Each polling district has exactly one polling station. The source of the geojson file can be found in [9].

The planning process for the municipal council and district council elections in October 2020 started in June and at this time, the further course of the COVID-19 pandemic was not clear. While the number of cases was rather low in June, the actual number was hard to predict, and a higher number of cases was to be expected for October. Furthermore, at that time, COVID-19 tests were not yet available for everyone, but rather only for those persons with symptoms. Therefore, simply testing the persons entitled to vote before they enter the polling booth, as we would do it now with the “green pass”, was not possible. To be prepared for every scenario, different hygiene measures were developed in collaboration with hygiene experts, as elections usually would bear an infection risk. These measures could come into effect depending on the number of COVID-19 cases active during the election. These concepts affected every task during the election, including the training of election personnel, the setup of polling stations, and the counting process. To minimize the risk of election-caused COVID-19 clusters, external measures such as more dates for training, more allocated time for the setup and more rooms and personnel for the counting were applied. It was agreed that these measures would suffice for these contexts.

The assessment of measures of the actual voting process is more complex, as it not only involves the voting personnel, which can be trained, but also the voters, whose number is not known in advance. While the adherence to the measures within polling stations can be monitored by the staff, a problem lies in the emerging queues.

Most hygiene measures that were developed for everyday life (like providing hand sanitizer, keeping distance and wearing a face mask) could be applied to the election too. Furthermore, the election workers didn’t only wear a face mask, but also a face shield. However, each voter needs to be identified before casting the vote, and this contradicts with wearing a face mask during the whole process. In order to identify the voters without putting anyone at risk to be infected with COVID-19, two possible hygiene strategies were defined (see also [10]):

- <u>Strategy A:</u> Each voter is asked to maintain a safety distance to the election workers, take off their face mask in order to be identified, and to put it right back on afterwards.
- <u>Strategy B:</u> Each voter is asked to go into a perspex booth, take off the mask there, wait to be identified, put the mask back on and leave the perspex booth.

It goes without saying that hygiene strategy B is safer than A, but also takes more time.

Clearly, both of the defined hygiene strategies might cause long queues in front of the polling station, as due to them, the voting process will take longer in total. Counter measures could be applied, if potential bottlenecks are known upfront.

If only few problematic polling stations were to be identified, one could think about applying additional measures to these stations directly, like offering an additional polling booth. Also merging or splitting of voting districts is possible.

Therefore, modeling and simulating the voting process and estimating the length of the queue for each voting district before the actual election takes place was deemed to be of great importance.

### Boundary conditions and comparison

Answering the key question of the planning process, namely how to avoid the development of long waiting queues in front of the polling stations, with the help of a simulation model requires to define the observed system’s borders — a task which in this case proves quite challenging. In principle, there are two possibilities to tackle the problem: First, the task can be analyzed from a *microscopic* point of view regarding all processes within one single polling station. In this process, we need to analyze and reduce the risk of infection in a single building. Second, the problem can be regarded on a *macroscopic* level, considering the election process in the whole city and all interactions involved.

Clearly, the microscopic point of view would neglect the pandemic nature of the phenomenon. Since these elections are taking place in an urban area with a heterogeneous population (in terms of population density as well as demographic features – which means different infection risks) at several hundred polling stations, the macroscopic perspective is hard to neglect.

As the election takes place in a very limited time-frame only, i.e. the 10 hours during which the polling stations are open, it was possible to decouple these perspectives and we subsequently split the problem into two: the *socio-demographic aspect* which assesses the general epidemic situation for the Vienna metropolitan area and the *logistical aspect* which deals with the local risks of infection at the polling stations.

With the former being not only harder to control (until today, policy makers are struggling with getting a grip on the epidemic) but also much slower to affect, we chose to incorporate it as a *“prognosis of boundary conditions”* within which the logistic part of the problem had to be solved.

Modeling and analyzing the queuing times for the election, the natural choice of modeling approach is a discrete event based queuing system, where the voters are the entities that run through a system of servers and queues that represent the different stations during the voting process. While a more detailed model using agents and their individual (locally optimal) paths would have been possible (see for example [11, 12] or also [13, 14]), an agent-based approach was deemed too detailed because of the sequential nature of the voting system, as there is no prioritization within the waiting voters and always the first in the queue is the next to vote (i.e. there is no overtaking, and therefore the individual paths of the voters are not that important).

The queuing system is typically analyzed either with classical steady state analysis or with stochastic simulation. However, considering steady state analysis, the system doesn’t fulfill all conditions, that are required to yield reasonable results (see also discussion). Therefore, the system is analyzed using stochastic simulation.

### Discrete event model

The following section deals with the implementation of the discrete event model. First, an overview over the model is provided. Next, the implemented scenarios and the calibration process are described. Finally, a dashboard is shortly introduced which visualizes the simulation results and helps with the decision making process.

### Simulation Model & Implementation

Our discrete event simulation model (DES) can be considered as the interface between epidemiology, which is the reason for our research, and logistic, which is the solution. In the following, we give a short introduction into the modeling process and conclude with a reproducible model definition and some important details about its implementation.

### Process Description and Modelling

Considering the different strategies of hygiene measures and assessing their impact on waiting times and queues, it was necessary to extend the established models. While the hygiene measures A (taking off the mask for identification with safety distance) and B (taking of the mask for identification in a perspex booth) mainly affect the service times for the different voting processes, the need of social distancing and the limitation of voters allowed inside the polling station at any given time revealed a new challenge. After entering the polling room, the voters must identify themselves at the electoral commission and collect their ballot card. They then proceed to the polling booth. Thereafter, they deposit their ballot card at the electoral commission before leaving the polling room. Since the overall process essentially consists of different points in the building where people queue in line, the simulation model was defined as classic tandem server-queue model.

As most polling stations are located in schools or other public buildings and the used rooms often only have one entrance/exit, where voters can get in and out, they block each other while entering and exiting. Additionally, while under normal circumstances it would be possible for voters to deposit their ballot card while another voter is identified at the commission, due to distancing rules voters have to wait at the entrance until one of the previous voters leaves the room and may have to wait for the completion of the identifying process of another voter until they can hand over their ballot card. These facts were taken into account by adding another two server-queues to the model that represent the entrance and exit of the building. Since the servers use joint resources, the entrance and the exit, and the electoral commission and the ballot box are linked to one joint resource pool, respectively.

Moreover, different polling stations show different properties due to the different demographic and socioeconomic features of the voting population. First, elderly persons are often linked to reduced mobility. Hence their voting process takes more time. Secondly, voter preparedness is typically different in different regions. For example, if a voter brings their voting information, they are found faster in the system and therefore the identification is faster. Third, in some regions voters are more likely to use the option of preferential voting, which takes more time. To depict these features, the tandem server queue is designed to use parametric entities.

### Model definition

The model is designed as a classic DES model using parametric entities. The entities represent voters and are assigned the following three boolean-valued attributes and an age:

- *LM* : *true*, iff the represented voter has limited mobility;
- *V I*: *true*, iff the represented voter brings their voting information;
- *PV* : *true*, iff the represented voter uses preferential voting.
- *AGE*: The age of the voter, depending on the age distribution of the polling station’s district.

Furthermore, we regard the following properties of the overall polling station:

- *N* : Total number of people entitled to vote in this polling station.
- *age*: The age distribution in the polling station’s district.
- *prob*: The probability distribution, by which time voters arrive at the polling station.
- *HALL*: *true*, iff the polling station is specified as hall-type in contrast to room-type polling stations. Hall-type polling stations present longer walking distances within the station.
- *n*: The number of polling booths: Generally, two if *N <* 1000, three otherwise.

The first three properties are used for initialization of the entities, the latter two influence the speed of the voting process. Furthermore, the service times depend on which hygiene scenario is applied, either none, SA (Scenario A, no perspex booth) or SB (Scenario B, with perspex booth).

In Figure 2 the tandem server queue representing the voting process is shown. The model consists of the five servers, *Entrance* (E1) and (E2), *Electoral Commission* (C1) and (C2), and *Voting Booths* (B = *{* B1, …, Bn*}*). Server B is a standard multi-server with one queue (QB). Server C1 and C2 are single servers that share the same resource RC. To ensure that voters are not blocked on their way out, C2 has prioritized access to RC compared to C1. That means, that waiting entities in QC1 are only served, if QC2 is empty.

**Fig 2.**
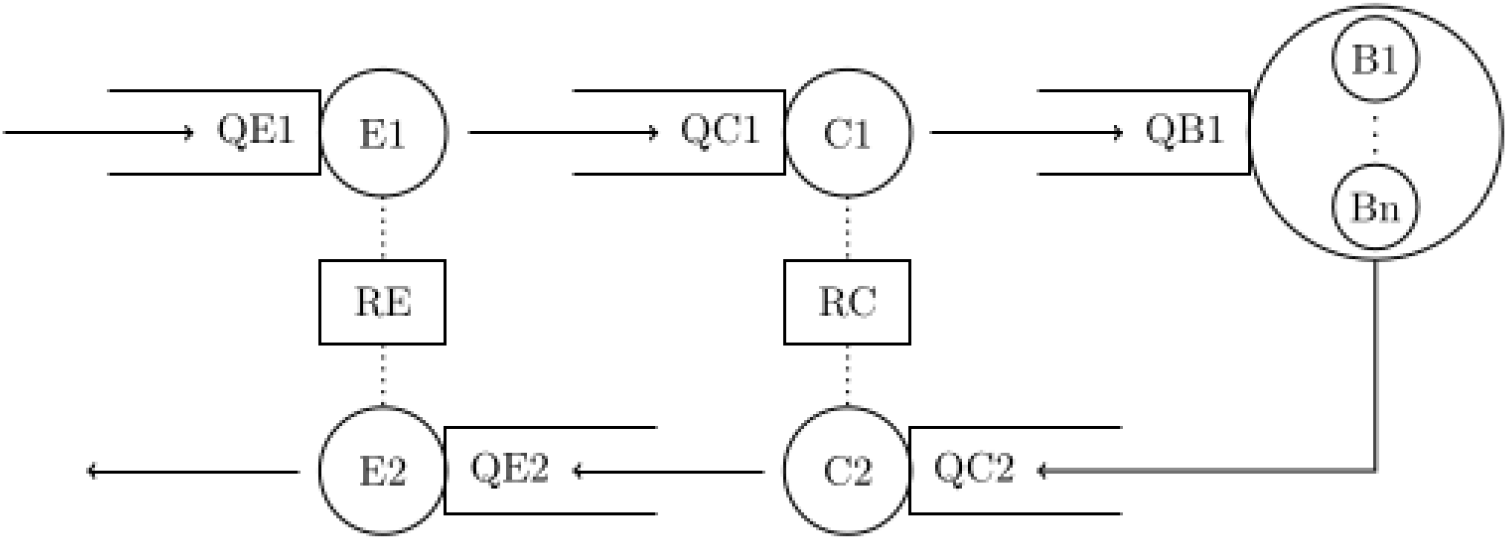
Depiction of the tandem server queue model for voter flow with servers Entrance (E1 and E2), Electoral Commission (C1 and C2), and Voting Booths (B1, …, Bn), their respective queues, and the two shared resource pools RE and RC.

Servers E1 and E2 also share the same resource RE whereas E2 is prioritized. Server E1 has two additional guard functions: Entities queuing in QE1 are only served if (a) the total number of entities in the system except from queue QE1 itself, is smaller than a fixed upper limit, and (b) the resource RC is idle.

All five queues are designed as FIFO queues without capacity limit.

The service times of the five servers are stochastic, and can be written as *µ* + *X*, whereas *X* ∼ *Exp*(*λ*). Here, *λ* (which we specify as the mean value of the exponential distribution) depends on the server, while *µ* depends on the server, properties of the processed entity, and specific properties of the polling station. All service times and their dependence on different properties are summarized in Table 1. *µ* calculates strictly from left to right (no order of operations), with base value *µ*_*base*_. The rightmost column shows the mean value parameter *λ* of the specific server.

**Table 1.** Service times of all five servers in the model

| Server | $\mu_{base}$ | *HALL* | *LM* | $\neg VI$ | *PV* | *SA* | *SB* | $\lambda$ |
| --- | --- | --- | --- | --- | --- | --- | --- | --- |
| E1 | 3s | +2s | $\times 2$ | | | | | 2s |
| C1 | 10s |  |  | +15s |  | +7s | +10s | 8s |
| B1 ... Bn | 30s |  | +15s |  | +30s |  |  | 30s |
| C2 | 3s |  | +5s |  |  |  |  | 2s |
| E2 | 3s | +2s | $\times 2$ | | | | | 2s |

### Initialization

All *N* entities arrive at QE1 based on upfront calculated arrival times. These are drawn randomly from the distribution *prob*. The distribution function is considered piecewise constant, Table 2 presents the probabilities per hour. As soon as an entity arrives in the queue, it is assigned a random age, based on the given age distribution *age*. Furthermore, the properties *LM, V I* and *PV* are specified with certain probabilities:

**Table 2.** Probabilities *prob* of voter arrival per hour

|  |  |  |  |  |  |
| --- | --- | --- | --- | --- | --- |
| hour | 7-8 | 8-9 | 9-10 | 10-11 | 11-12 |
| p | 0.06682 | 0.06682 | 0.06682 | 0.13471 | 0.13471 |
| hour | 12-13 | 13-14 | 14-15 | 15-16 | 16-17 |
| p | 0.13075 | 0.13075 | 0.07976 | 0.07976 | 0.10910 |

- *P* (*LM* = *true*|*AGE* < 65) = 0.136,
- *P* (*LM* = *true*|*AGE* ≥ 65) = 0.336,
- *P* (*V I* = *true*) = 0.67,
- *P* (*PV* = *true*) = 0.1.

### Implementation

The model was implemented in a self-developed, Java-based framework for agent-based and discrete event models called ABT [15].

### Calibration and validation

As encouraged in the literature, and in order to derive useful recommendations for actions, we made substantial efforts to calibrate the model as well as possible, i.e. to set the parameters reasonably.

The parameters for the arrival times of the voters are chosen according to the arrival times of past elections [16, 17]. The age distribution for the district of each polling station is drawn from statistical data [18, 19]. The amount of voters bringing their voting information and using preferential voting corresponds to 67% and 10%, respectively, according to the experts from the Municipal Department 62, which is responsible for elections.

The times each voter needs at each place in a polling station (to walk to the electoral commission, to be identified, to vote) are drawn from an exponential distribution. The parameters of these distributions were initially estimated by election workers of past elections. Afterwards, these estimations were calibrated assuming a voter turnout of 75% with 250 000 postal voters, voters with limited mobility according to prevalence and no hygiene strategy. This scenario corresponds approximately to the conditions during the last election in Austria, which was the national council election in 2019. Within this scenario, the parameters were carefully adjusted using a bisection algorithm, until the simulation yielded reasonable waiting queues. However, as data from queues have not been measured so far, “reasonable waiting queues” were in fact hard to quantify; the Municipal Department 62 could not provide any data on this, either. But due to their experience, they stated that the longest queue forming in front of a polling station during the election day never exceeded 15 persons. Therefore, we looked for a parametrization of the model such that the longest queue was lower or equal to 15 in every polling station.

Via the bisection algorithm, we found a calibration which yielded maximum queues of 15 people or less in 1439 of the 1456 voting districts (i.e., 98.70%). In the remaining 17 stations, however, extensive queues of up to 132 persons would emerge, which clearly made a closer inspection of these stations with the Municipal Department 62 necessary. This inspection showed that these stations were already known as potential risk stations and were thus under observation. These stations, for example, belong to districts with care homes; therefore, the average age is comparatively high, and the voters need accordingly long to vote in the simulation. In reality, care homes offer almost exclusively postal voting, which is why these stations are actually no risk stations. The other stations belong to known big voting districts, and were already planned to be split up into two smaller voting districts. As these stations were already under observation, we excluded them from the further analysis.

Via careful inspection from the planning experts using the implemented dashboard (see below), the model passed general face validity tests. Since the model properly displayed already known bottlenecks of a standard election process without additional hygiene measures, the simulation model achieved a certain level of quantitative validity as well. Note, that there is no data about queue lengths of previous elections available that would allow a more detailed validation process.

### Scenarios

The model allows the simulation of several scenarios and strategies. First, the amount of voters and the number of postal voters were varied, as well as the amount of people with limited mobility and the number of additional polling booths. Additionally, the different hygiene strategies were simulated. Table 3 provides an overview over the varied values.

**Table 3.** Scenarios covered by our simulation

| Parameter | Values |
| --- | --- |
| Voter turnout | 65%, 70%, 75%, 80% |
| Number of postal votes | 250 000, 300 000, 350 000, 400 000, 450 000, 500 000 |
| Voters with limited mobility | According to prevalence, under prevalence, above prevalence |
| Hygiene Strategy | No strategy, strategy A, strategy B |
| Additional polling booths | 0, 1, 2 |

To smooth statistical effects, the mean of 5 simulation runs was considered. All possible parameter combinations have been simulated. However, in order to demonstrate the impact of the single parameters on the queues, only one parameter has been varied at a time, to create the plot of Figure 3. Based on the above mentioned baseline scenario, each of the parameters has been varied, and the longest queue, that appears throughout the day in one of the polling stations in each district, has been calculated. Considering these results, the following observations can be made:

**Fig 3.**
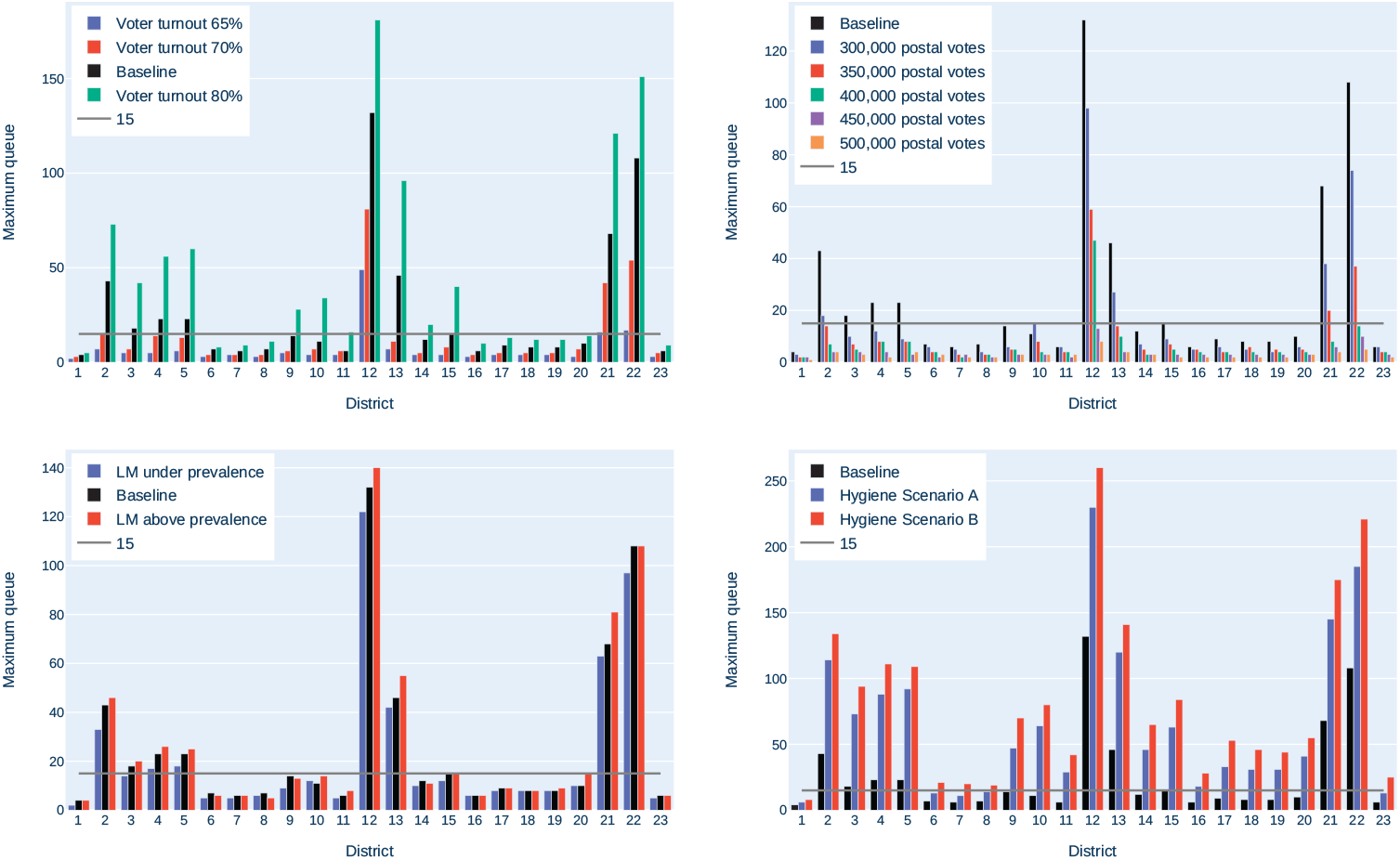
Impact of different voter turnouts, postal voters, limited mobility (LM) and hygiene scenarios on the maximum queue in each district in Vienna. The baseline uses a voter turnout of 75%, 250 000 postal votes, limited mobility according to prevalence and no additional hygienic measures.

- The number of people appearing at the polling stations has a crucial impact on the queues. From a democratic point of view, a high voter turnout is generally desirable, but the number of postal voters can be increased. For example, having 300 000 instead of 250 000 postal voters reduces the maximum emerging queue in some districts by half. Thus, using postal votes should be further encouraged.
- The amount of voters with limited mobility has hardly any impact on the queue lengths. Therefore, in the following analysis, only “limited mobility according to prevalence” will be considered.
- The difference from “no hygiene strategy” to “hygiene strategy A” is larger than the difference from “hygiene strategy A” to “hygiene strategy B”. Seeing this, hygiene strategy B should be preferred, as it is safer without having that much impact on the waiting queues.
- It happens, when comparing two scenarios, that the maximum queue is longer in a scenario, where a shorter queue would have been excepted, or vice versa. For example, considering the maximum queues for different values of limited mobility, the maximum queues are not necessarily increasing with increasing limited mobility. This happens especially in smaller districts, due to random fluctuation.

### Dashboard

In order to visualize the scenarios and thus, to improve the decision making process, a dashboard containing the simulation results was implemented. In this dashboard, it is possible to set a scenario according to the values shown in Table 3. As measure of interest, for each scenario and for each district, the longest queue forming at one point of the election day in front of the polling station was chosen. These values are visualized in a choropleth map of Vienna, in order to identify polling stations with a need for action, see also Figure 4.

**Fig 4.**
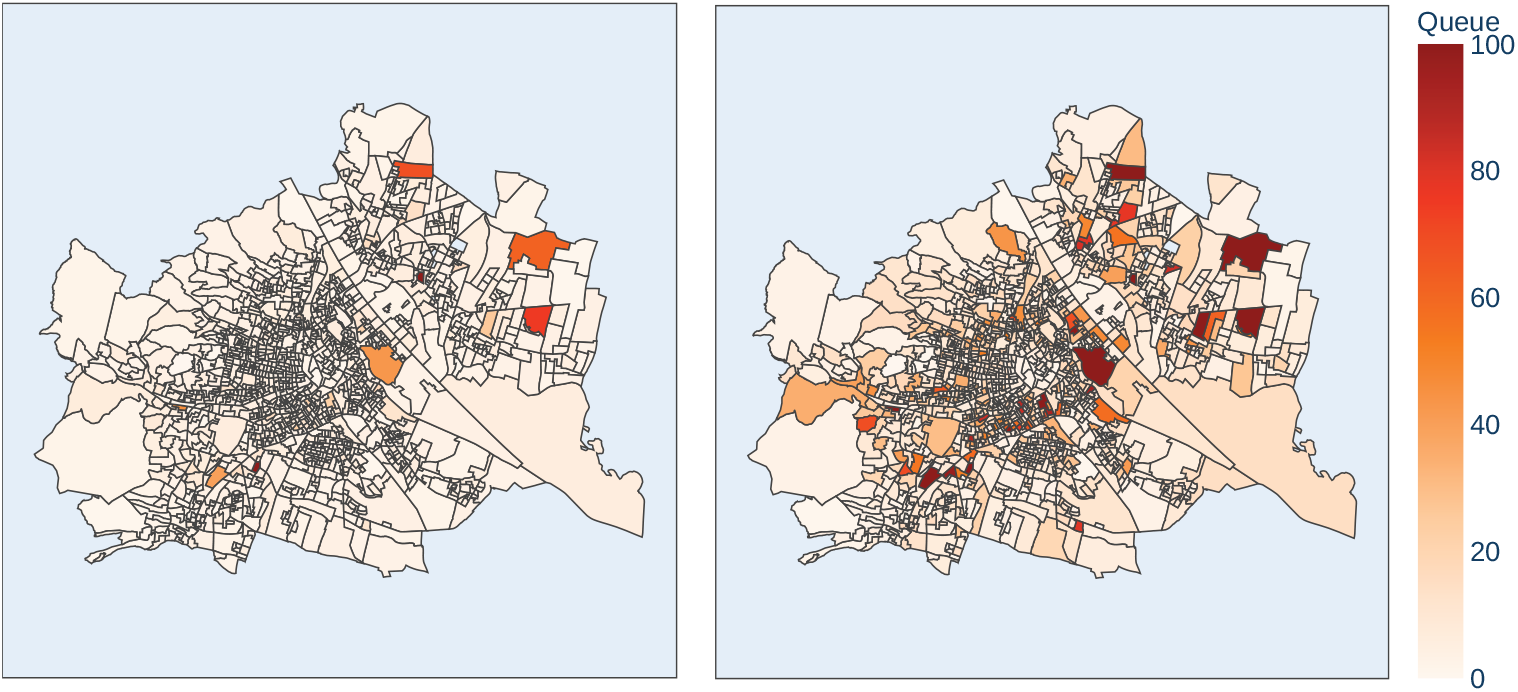
Simulated waiting queues by the model. Left: Baseline Scenario (17 risk stations), Right: Hygiene strategy B added to baseline (351 risk stations). For the sake of visibility, the darkest color was chosen for all districts that have a maximum queue of 100 persons or more. The source of the geojson file can be found in [9].

## Results

Considering the goal of the analysis, there are two kinds of results that can be considered: queue lengths, meaning both simulated results from our model and reported waiting queues at the election day, and SARS-CoV-2 infection numbers, measured a few days after the election.

### Model results

Together with the Municipal Department 62, the following baseline election scenario was defined for the upcoming election: A voter turnout of 70%, 300 000 postal votes, voters with limited mobility according to prevalence and hygiene strategy B. Based on this election scenario, the voter turnout and the amount of postal voters has again been varied, as can be seen in Figure 5. It still holds for this scenario, that the maximum queues are crucially influenced by these parameters, and that postal voting should be encouraged.

**Fig 5.**
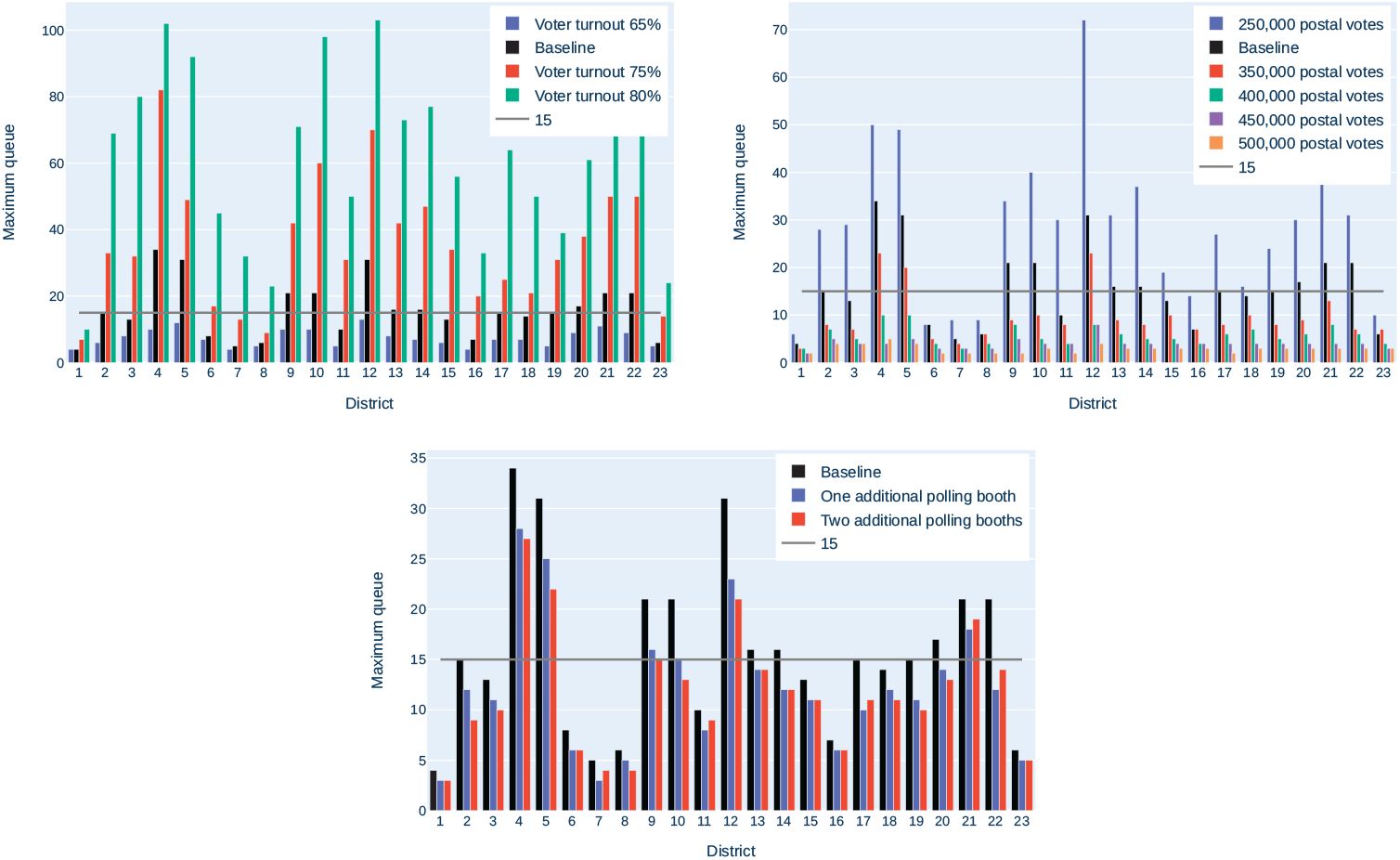
Impact of different voter turnouts, postal voters and additional polling booths on the length of the maximal queue in each district in Vienna, neglecting the already known risk stations.

For these assumptions, our model identified 41 polling stations as potential risk stations, meaning that in front of these stations queues of 15 people or more could emerge. 17 of these stations (actually almost the top 17), however, correspond to the already mentioned known stations that are treated separately. Therefore, these stations are excluded from the following investigations.

This left 24 new potential risk stations, all of which have comparatively many people entitled to vote. Based on this, two new simulations were run, with one and two additional polling booths in each polling station, respectively. The simulation runs showed, that additional voting booths in these stations don’t alleviate the queues sufficiently (i.e. maximum queues of 15 persons or less), as it can also be seen in Figure 5. This can be explained with the actual bottleneck of the election being the entrance, not the voting process itself. At the entrance sits the election commission. This is a group of local politicians and volunteers, that look over the whole election process. At a time, only one voter can enter the voting station, as they keep all members of the election commission busy. Plus, entering is the only part of the election whose duration is extended due to the hygienic measures, as they lead to prolonged ID checking. Two additional voting boxes hardly had any impact.

Splitting up a polling station into two stations, on the other hand, results in less people being entitled to vote in the respective station, and thus, in shorter waiting queues. Therefore, the Municipal Department 62 decided to split up some of the risk stations. In total 32 polling stations were split up into two stations.

### Actual election participation results

Unfortunately, our attempts to gather queue length data for proper reevaluation of the model results were not successful for different administrative and data privacy reasons.

The complete absence of negative news about crowded polling booths – in particular in times in which almost any mass gathering of persons causes a COVID-19–news headliner – suggests that there were actually no gatherings. However, of course this does not prove that there were no gatherings, nor whether it was actually the combination of the strategies that has successfully prevented crowding.

Of the 1 362 789 people that were eligible to vote, 786 777 people cast their vote, yielding a voting turnout of 57.73%. The turnout is significantly lower than in 2015, having a turnout of 65.79%. This might not only be explained by COVID-19, but also by some domestic affairs. Of all voters, 339 445 (43.14%) used postal votes. This is a comparably high number, as in 2015, only 159 226 voters (12.00%) used postal voting [8].

### Reported COVID-19 case numbers

Still, 447 332 persons voted in person, bearing an increased infection risk for the election day. Indeed, infection numbers in Austria strongly increased by the middle of October, which coincides with the days after the election. However, a closer inspection of the incidences in the single federal states shows, that the incidences in Vienna were comparably low; actually, incidences in Vienna were the only descending ones in Austria, see also Figure 6. While the election can not be held responsible for the decreasing numbers in Vienna, it can at least be stated, that it has not contributed to an increase of incidences. [1] In order to fully evaluate the impact of the election on the incidences, a more thorough analysis would be required, e.g. like it was done in [20]. This kind of analysis, however, was beyond the scope of this work.

**Fig 6.**
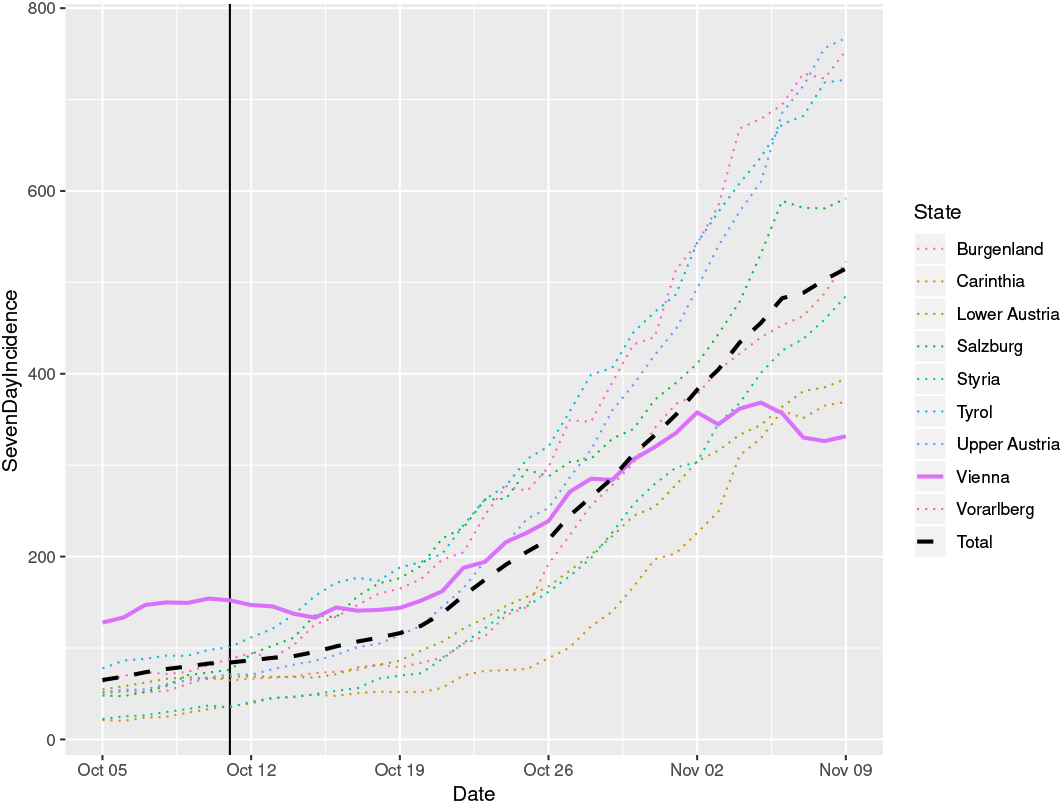
Incidences in Austria and in each federal state before and after the election date according to official data [1]. The vertical line denotes the election day.

## Discussion and conclusion

Our work shows that the usage of our discrete event simulation model can help to point out polling stations where long queues in urban elections could develop. In combination with proper hygiene concepts, it contributes to allow safe elections even in times of epidemics. However, it is important not to underestimate the risks that occur from the pandemic, and to take appropriate precautions.

The decision to model the voting process as a queuing model corresponds to the findings of the available literature on this topic. The question of how to model the voting process and analyze the queue lengths during this process was predominantly investigated in the US [21–29] but also in other countries like Nigeria [30] or Hong Kong [31]. Most of these papers, especially the ones from the US, focus on the optimal distribution of voting machines. This is due to the widespread use of such machines in that country. Hence, the scarcity of these resources presents the biggest bottleneck during the US voting process. Additionally, different waiting times in different voting precincts are a major problem. Other papers [28, 30, 31] are more concerned on optimizing the actual voting process by providing more resources such as voting booths or identification desks. Big differences between the models are mainly due to varying voting processes, for example in Nigeria the overall process is split into two phases, where the voters have to get accredited in the morning and the actual vote is performed in the afternoon. Technologically, the queuing system is typically analyzed in one of two ways. The first approach is classical steady state analysis [24, 28], but using steady state analysis creates a number of problems and inaccuracies for the analysis:

- Using steady state analysis requires the system to be in a steady state. Due to the relatively short voting time, it is not always possible to ensure that the system even reaches this state.
- If the arrival rate is greater than the servicing rate of the system, the queues become unbounded, and therefore a steady state can never be reached.
- Queuing systems base their analysis on arrival rates and service times that are randomly distributed, but the underlying distribution cannot change over time. This restriction is particularly limiting for the consideration of voting processes, as the arrival rates are typically dependent on the time of day, and the actual voting times depend heavily on the type of voter. For example, disabilities lead to higher voting times, but also voters that use preferential voting take longer than voters who only vote for a political party.

These problems show that steady state analysis is not a feasible method for studying voting processes, which is acknowledged in most of the literature. The second approach for analyzing queuing systems is stochastic simulation, which is also the preferred method in most of literature [21, 23, 25–27, 30, 31]. This ansatz enables not only a more detailed stochastic analysis, but also the possibility to vary parameters over time and describe voters with different time requirements. Basically all of the presented papers agree that carefully estimated arrival rates that can vary throughout the day and servicing times for the different processes, are key for a sophisticated prediction of waiting times and queue lengths.

However, the limitations of our study need to be stated. For our simulation model, the following simplifications had to be made:

- No reneging: Voters don’t leave the queue if the wait is too long. This could limit the application to other elections where queues are typically longer.
- No precise approximation of persons with limited mobility in each district: Our estimation of the amount of people with limited mobility relies solely on the age distribution in this district.
- No realistic distribution of postal voters: Our model assumes that the amount of postal voters is the same across all age groups. This is, for example, not true for people living in care homes, as they use postal voting almost exclusively. This yielded unreliable results in districts with large care homes. Furthermore, the amount of postal votes depends on the socioeconomic population structure and varies within the districts. And finally, it is possible to order postal voting and nevertheless enter a polling station in order to deposit the vote. This is done, for example, when a voter wants to vote in a different polling district than their home district. This wasn’t considered either by the model.

Additionally, we had limited data for both parametrization and validation. Queue-data have not been measured and collected so far, and thus, the calibration process could only be validated visually, instead of quantitatively. As this work focused on identifying those stations where long queues could emerge, instead of on the actual queue lengths of the single stations, the visual validation was sufficient. However, if a more precise study with reliable queue length numbers is required, more data would be necessary to parametrize and validate the model correctly.

In general we showed that modeling and simulation is a valid way to test new conditions in an election. In this case, the precautions that were taken in order to avoid a new spread of COVID, lead to an unavoidable prolongation in the voting process. The simulation showed, however, that the process was not prolonged in a way such that long queues formed in front of the polling stations. The actual election participation results further showed that the effort to motivate people to vote postally was successful, which also contributed to both short queues and short waiting times. Nevertheless, the model can easily be adapted to simulate any other changes in the voting scheme as well.

Altogether the established strategy resulted in an election which can be considered as “smooth”. It should however be taken into account that this is usually the case in Austria. It rarely happens that queues in front of polling stations form at all, and if, these are mostly short ones. Still, it is worth acknowledging that this condition was threatened by the additional measures, but could be upheld.

Finally, the analysis of the reported case numbers in the city of Vienna indicated that the election did not cause a temporary upswing of the epidemic. Surely, there are many factors that might have prevented additional infections such as wearing face-mask, large and well ventilated polling rooms, keeping the distance in the queues, low voter turnout, campaigns for mail voting, or simply because voting is a quick process that does not require talking. Nevertheless, it is legit to assume that the preventive measures against queuing bottlenecks that were detected by the simulation model contributed to avoid election related infection clusters – and this was, essentially, the uppermost goal of the simulation study.

## Data Availability

All data are fully available without restriction.
The data underlying the results presented in the study are available from

https://git.dwh.at/nweibrecht/SimulatedQueues

## Acknowledgments

We thank the Austrian National Public Health Institute (Gesundheit Österreich GmbH, GÖG) for their help in discussing and interpreting the COVID-19 incidences.

## References

1. AGES Dashboard COVID19 [Dashboard];. Available from: https://covid19-dashboard.ages.at/dashboard.html.

2. Warteschlangen vor den Wahllokalen. Wiener Zeitung. 2017 Oct;Available from: https://www.wienerzeitung.at/nachrichten/wahlen/nationalratswahl-2019/nationalratswahl-2017/923105-Warteschlangen-vor-den-Wahllokalen.html.

3. Hötzendorfer H, Popper N, Breitenecker F. Temporal and Spatial Evolution of a SIR-type Epidemic – ARGESIM Comparison C17 - Definition. Simulation News Europe SNE. 2004;(41):3.

4. Miksch F, Jahn B, Espinosa KJ, Chhatwal J, Siebert U, Popper N. Why should we apply ABM for decision analysis for infectious diseases?—An example for dengue interventions. PLOS ONE. 2019 Aug;14(8):e0221564. Available from: https://dx.plos.org/10.1371/journal.pone.0221564.

5. Bicher M, Rippinger C, Urach C, Brunmeir D, Siebert U, Popper N. Evaluation of Contact-Tracing Policies against the Spread of SARS-CoV-2 in Austria: An Agent-Based Simulation. Medical Decision Making. 2021 May;p. 0272989X2110133.

6. Rippinger C, Bicher M, Urach C, Brunmeir D, Weibrecht N, Zauner G, et al. Evaluation of undetected cases during the COVID-19 epidemic in Austria. BMC Infectious Diseases. 2021 Jan;21(1):70.

7. Bicher M, Zuba M, Rainer L, Bachner F, Rippinger C, Ostermann H, et al. Supporting Austria through the COVID-19 Epidemics with a Forecast-Based Early Warning System. Health Policy; 2020. Available from: http://medrxiv.org/lookup/doi/10.1101/2020.10.18.20214767.

8. Gemeinderatswahlen 2020;. Available from: https://www.wien.gv.at/wahlergebnis/de/GR201/index.html.

9. Katalog WFS GetFeature (JSON) - Gemeinderatswahl 2020;. Available from: https://www.data.gv.at/katalog/dataset/stadt-wien_wahleninwienwahlsprengel/resource/8eff5961-9359-4bb7-9453-974f4248fd71.

10. Wien-Wahl 2020: Sicher wählen am Wahltag;. Available from: https://www.wien.gv.at/presse/2020/10/02/wien-wahl-2020-sicher-waehlen-am-wahltag.

11. Tauböck S, Breitenecker F, Wiegand D, Popper N. The <morespace> Project: Modelling and Simulation of Room Management and Schedule Planning at University by Combining DEVS and Agent-Based Approaches. Simulation News Europe SNE. 2011 Apr;21(1):11–20.

12. Bruckner M, Tauböck S, Popper N, Emrich S, Roszenich B, Alkilani S. Modelling and Simulation of Student Pedestrian Traffic at University Campus. Simulation News Europe SNE. 2012 Aug;22(2):95–100.

13. Glock B, Wurzer G, Breitenecker F, Popper N. Reverse Engineering Hospital Processes Out of Visited Nodes. In: Al-Begain K, Al Dabass D, Orsoni A, Cant R, Zobel R, editors. EUROSIM 2013 8th EUROSIM Congress on Modelling and Simulation. Cardiff: EUROSIM 2013 8th EUROSIM Congress on Modelling and Simulation; 2013. p. 312–317.

14. Wurzer G, Lorenz WE, Rößler M, Hafner I, Glock B, Bruckner M, et al. MODYPLAN: Early-Stage Hospital Simulation Based on Treatment Chains. IFAC-PapersOnLine. 2015 Jan;48(1):868–873.

15. The Power of the ABT Simulation Framework;. Available from: https://www.dwh.at/en/blog/the-power-of-the-abt-simulation-framework/.

16. Detailergebnisse der Geimenderatswahl 2015;. Available from: https://www.wien.gv.at/wahl/NET/GR151/GR151-109.htm.

17. Detailergebnisse der Nationalratswahl 2017;. Available from: https://www.wien.gv.at/wahlergebnis/de/NR171/index.html.

18. Bevölkerung nach Altersgruppen, Geschlecht und Gemeindebezirken 2019.;. Available from: https://www.wien.gv.at/statistik/bevoelkerung/tabellen/bevoelkerung-alter-geschl-bez.html.

19. Ausgewählte Ergebnisse aus der Abgestimmten Erwerbsstatistik und der Arbeitsstättenzählung 2018 (Gebietsstand 2020);. Available from: https://www.statistik.at/wcm/idc/idcplg?IdcService=GET_PDF_FILE&RevisionSelectionMethod=LatestReleased&dDocName=079497.

20. Zeitoun JD, Faron M, Manternach s, Fourquet J, Lavielle M, lefevre j. Reciprocal association between participation to a national election and the epidemic spread of COVID-19 in France: nationwide observational and dynamic modeling study. Public and Global Health; 2020. Available from: http://medrxiv.org/lookup/doi/10.1101/2020.05.14.20090100.

21. Grant FH. Reducing Voter Waiting Time. Interfaces. 1980;10(5):19–25.

22. Belenky AS, Larson RC. To Queue or Not to Queue. OR/MS Today. 2006;33:30–34.

23. Edelstein WA. New Voting Systems for NY— Long Lines and High Cost. New Yorkers for verified Voting; 2006.

24. Allen T, Bernshteyn M. Mitigating Voter Waiting Times. CHANCE. 2006;19:25–34.

25. Yang M, Fry MJ, Kelton WD. Are All Voting Queues Created Equal? In: Proceedings of the 2009 Winter Simulation Conference (WSC); 2009. p. 3140–3149.

26. Yang M, Allen TT, Fry MJ, Kelton WD. The Call for Equity: Simulation Optimization Models to Minimize the Range of Waiting Times. IIE Transactions. 2013 Jul;45(7):781–795.

27. Yang M, Fry MJ, Kelton WD, Allen TT. Improving Voting Systems through Service-Operations Management. Production and Operations Management. 2014;23(7):1083–1097.

28. Stewart III C. Managing Polling Place Resources. Caltech/MIT Voting Technology Project; 2015.

29. Wang XJ, Yang M, Fry MJ. Efficiency and Equity Tradeoffs in Voting Machine Allocation Problems. Journal of the Operational Research Society. 2015 Aug;66(8):1363–1369.

30. Olabisi UO. Modeling and Analysis of the Queue Dynamics in the Nigerian Voting System. The Open Operational Research Journal. 2012 Nov;6(1):9–22.

31. Au CH, Xu Z, Wang L, Fung WSL. Establishing a Three-Step Model of Designing the Polling Stations for Shorter Queue and Smaller Waiting Time: A Case Study Using Computer Simulation. Journal of Information Technology Case and Application Research. 2017 Oct;19(4):225–245.

